# Clinical progression in alpha-synuclein positive LRRK2-PD and sporadic Parkinson’s disease: a longitudinal analysis

**DOI:** 10.64898/2026.03.19.26348792

**Authors:** Lucy A Morse, Seung Ho Choi, David-Erick Lafontant, Caroline Gochanour, Lana M Chahine, Kalpana M Merchant, Barbara Wendelberger, Tanya Simuni, The Parkinson’s Progression Markers Initiative

## Abstract

**Background:** LRRK2-Parkinson’s disease (LRRK2-PD) is biologically heterogeneous with approximately 30% lacking aggregated alpha synuclein (αSyn) in cerebrospinal fluid by seed amplification assay (SAA). Prior work has suggested slower progression in LRRK2-PD compared to sporadic PD (sPD).

**Objective:** We aimed to assess how LRRK2-PD with αSyn aggregates on SAA (S+ LRRK2-PD) compares to S+ sPD.

**Methods:** Data from the Parkinson’s Progression Markers Initiative were used to compare S+ LRRK2-PD and S+ sPD cohorts propensity score-matched on age, disease duration, sex and levodopa equivalent dose (N = 79 per cohort). Baseline clinical and biological features and 4-year longitudinal features were assessed.

**Results:** At baseline, S+ LRRK2-PD participants had lower motor scores and dopaminergic deficit. Among measures showing within group progression, longitudinal trajectories did not differ significantly between groups.

**Conclusions:** Longitudinal clinical progression of S+ LRRK2-PD and sPD in the PPMI study is similar despite differences in baseline features.

---

The majority of individuals with sporadic Parkinson’s Disease (sPD) have evidence of misfolded and aggregated alpha-synuclein (αSyn) on cerebrospinal fluid (CSF) αSyn seed amplification assay (SAA) (S+).^1, 2^ By contrast, approximately one third of participants with LRRK2-associated Parkinson’s disease (LRRK2-PD) are negative for αSyn on SAA (S-), consistent with neuropathological data demonstrating absence of Lewy bodies in 25-60% of LRRK2-PD brain tissue.^2–7^

Prior reports suggest that LRRK2-PD is associated with less non-motor disease burden and slower disease progression as compared to sPD.^8–10^ Importantly, recent data indicate that there are differences in clinical measures between LRRK2-PD S+ versus S- individuals: at baseline, S- LRRK2-PD participants have lower Movement Disorder Society Unified Parkinson’s Disease Rating Scale (MDS-UPDRS) Part II and III scores as compared to S+ LRRK2-PD, lesser degree of dopaminergic deficit on imaging, and have a trend towards slower motor and functional decline over time.^7, 11, 12^

Considering these findings, we aimed to compare clinical and biomarker characteristics between S+ sPD and S+ LRRK2-PD in the Parkinson’s Progression Markers Initiative (PPMI) study. We hypothesized that baseline characteristics and longitudinal progression would be similar in both groups, consistent with shared underlying proteinopathy.

## Methods

The PPMI study protocol, and methods for assessment of CSF biomarkers including αSyn SAA have been published previously.^1, 13–15^ The PPMI sample included 165 participants with LRRK2-PD (carriers of *LRRK2* G2019S, R1441C/G/H, N1437H, and I2020T as confirmed by the PPMI Genetic Coordination Core),and 950 participants with sPD for whom genotyping and αSyn SAA status were available. Exclusion criteria included S- status, MSA type- SAA, and inconclusive, and absence of at least one visit with levodopa equivalent dose (LED) > 0. Individuals with presence of a non-LRRK2 pathogenic variant were also excluded. Participants with fewer than one year of follow up for LRRK2-PD or fewer than three years of follow up for sPD were further excluded, initially yielding 96 LRRK2-PD and 284 sPD participants. Enrollment criteria for the PPMI genetic cohort were different from sPD cohort allowing for longer disease duration (up to 7 years) and use of dopaminergic therapy.^14, 16, 17^ To account for differences at enrollment between the two groups, a propensity score matching approach was employed where sPD participants were matched 1:1 to LRRK2-PD participants based on age, sex, disease duration and LED. Participants were aligned on their first treated visit – defined as ‘baseline’ for the purposes of this analysis. The matching process utilized a caliper width of 0.20 times the standard deviation of the logit of the propensity score with a sequential greedy nearest neighbor matching approach.

Differences in baseline characteristics between groups were assessed using the Wilcoxon rank sum test for continuous variables, and Chi-Square (or Fisher’s exact test when at least one expected cell count <5) for categorical measures. All baseline and longitudinal characteristics included in the analysis are presented in the results section and summarized in the accompanying tables.

Longitudinal analysis included clinical measures assessed at each annual visit. Medication OFF Part 3 scores were missing on a proportion of participants, and were therefore not included. Generalized linear mixed effects (LMM) models with random intercept and slope and unstructured working correlation structure were employed to model the linear trajectory of continuous outcomes by subgroup (sPD or LRRK2-PD) over 4 years, with adjustment for baseline value of the outcome and time-varying LED. For binary outcomes, only random intercepts were included, with the linear trajectory of the log odds being modeled. To assess the possibility of non-linear trajectories and differences by sex, tests for differential quadratic and group-sex-time effects were also conducted. Interaction and time effects from the models and associated Wald p-values are reported. Given the exploratory nature of this study, the significance threshold was set to 0.05 (two tailed).

Statistical analyses were performed using SAS v9.4 (SAS Institute Inc., Cary, NC, USA; sas.com; RRID:SCR_008567).

### Data Sharing

Data used in the preparation of this article were obtained on March 31, 2025 from the PPMI database (www.ppmi-info.org/access-data-specimens/download-data), RRID:SCR_006431. For up-to-date information on the study, visit www.ppmi-info.org. This analysis was conducted by the PPMI Statistics Core and used actual dates of activity for participants, a restricted data element not available to public users of PPMI data. Statistical analysis codes used to perform the analyses in this article are shared on Zenodo (10.5281/zenodo.16814373). Protocol information for PPMI Clinical - Establishing a Deeply Phenotyped PD Cohort AM 3.2. can be found on protocols.io or by following this link: https://dx.doi.org/10.17504/protocols.io.n92ldmw6ol5b/v2.

## Results

### Baseline characteristics

79 participants were included in the analysis per group after propensity matching. Participant selection and reasons for exclusion are presented in Supplementary Figure 1. Demographic and disease characteristics of the cohort prior to matching are presented in Supplementary Tables 1 and 2. Table 1 shows baseline demographic, clinical and biological characteristics of the matched groups. The majority of LRRK2-PD participants (96%) were G2019S carriers. At baseline, the S+ LRRK2-PD group had milder degree of functional impairment and motor dysfunction as indicated by higher scores on the Modified Schwab and England scale (median [IQR] 90 [90–100] versus 90 [80–90], p < 0.001), lower MDS-UPDRS part III ON scores (median [IQR] 16 [10–24] versus 21 [15–31], p = 0.011), lower Tremor score (ON) (median [IQR] 2 [0-5] versus 4 [1-6], p = 0.022), and lower Total MDS-UPDRS scores (median [IQR] 29 [20–42] versus 34 [27–51], p = 0.027) as compared to sPD. A lesser degree of dopaminergic dysfunction as characterized by lowest putamen ratio was observed in LRRK2-PD participants compared to sPD (median [IQR] 0.27 [0.24-0.37] versus 0.25 [0.20-0.31], p = 0.016). There were no differences between groups in fluid biomarkers including measures of CSF amyloid beta 1-42 (Aβ_1-42_), total tau, phospho-tau 181, or serum neurofilament light chain (NfL).

**Table 1.**
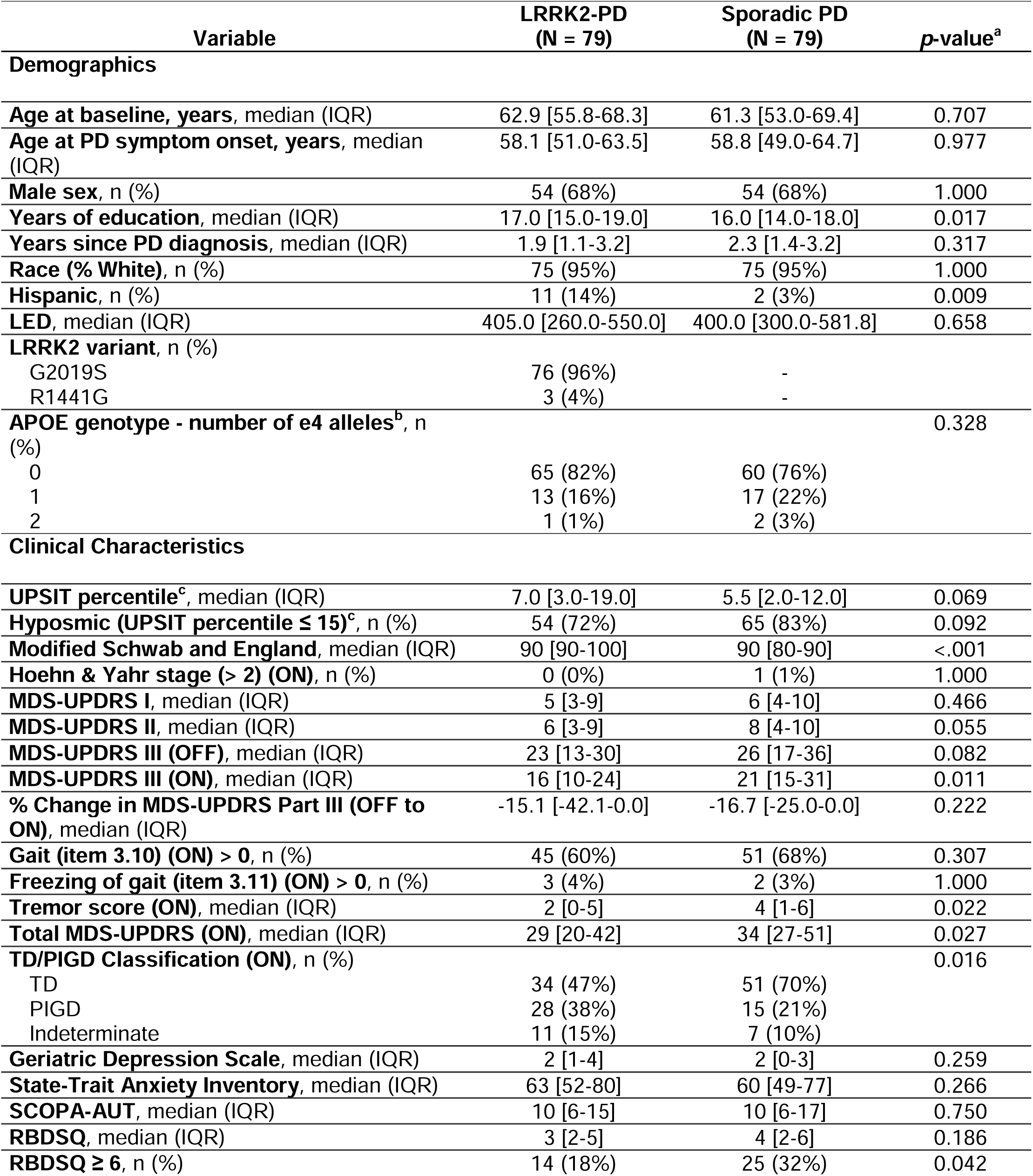
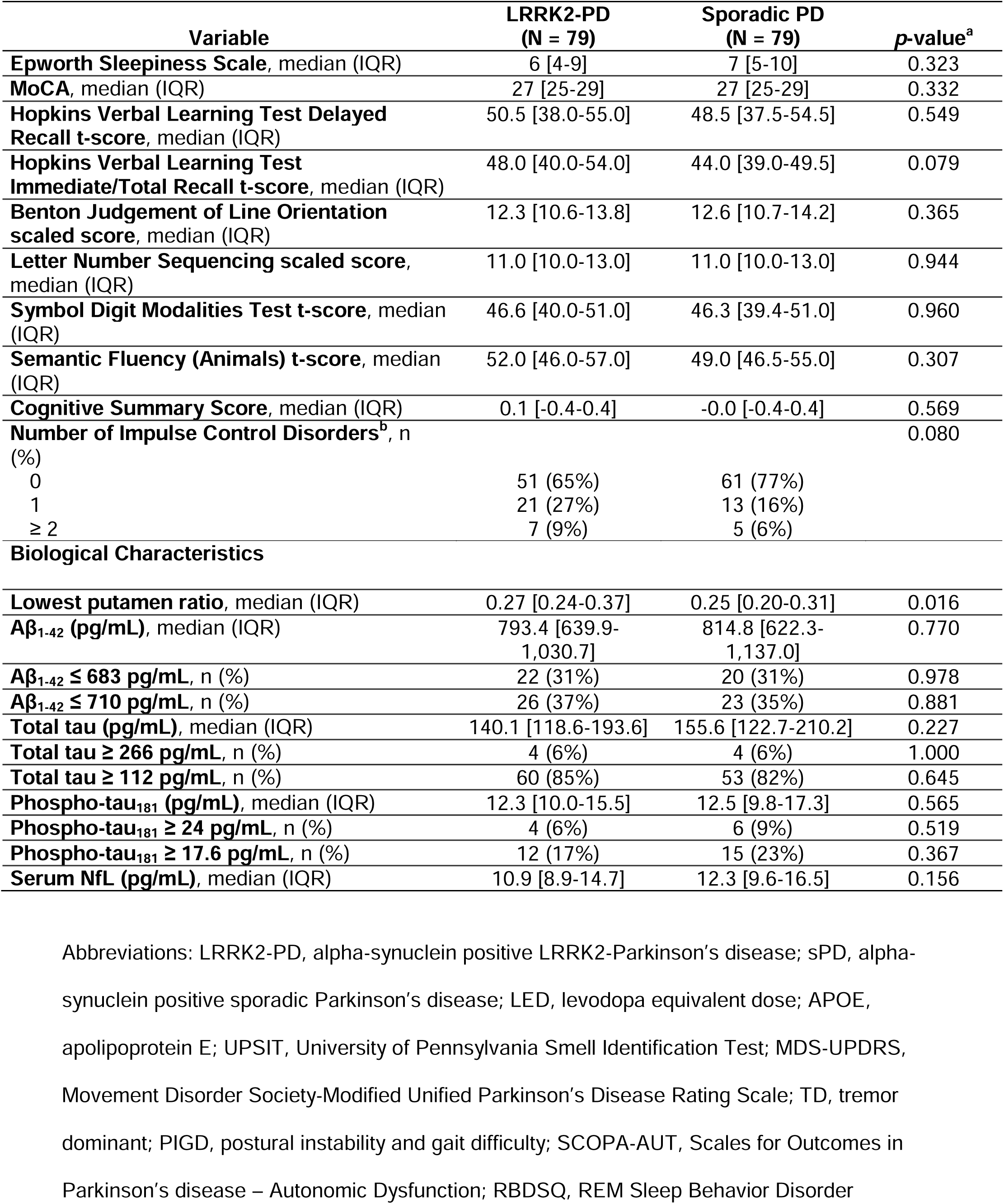
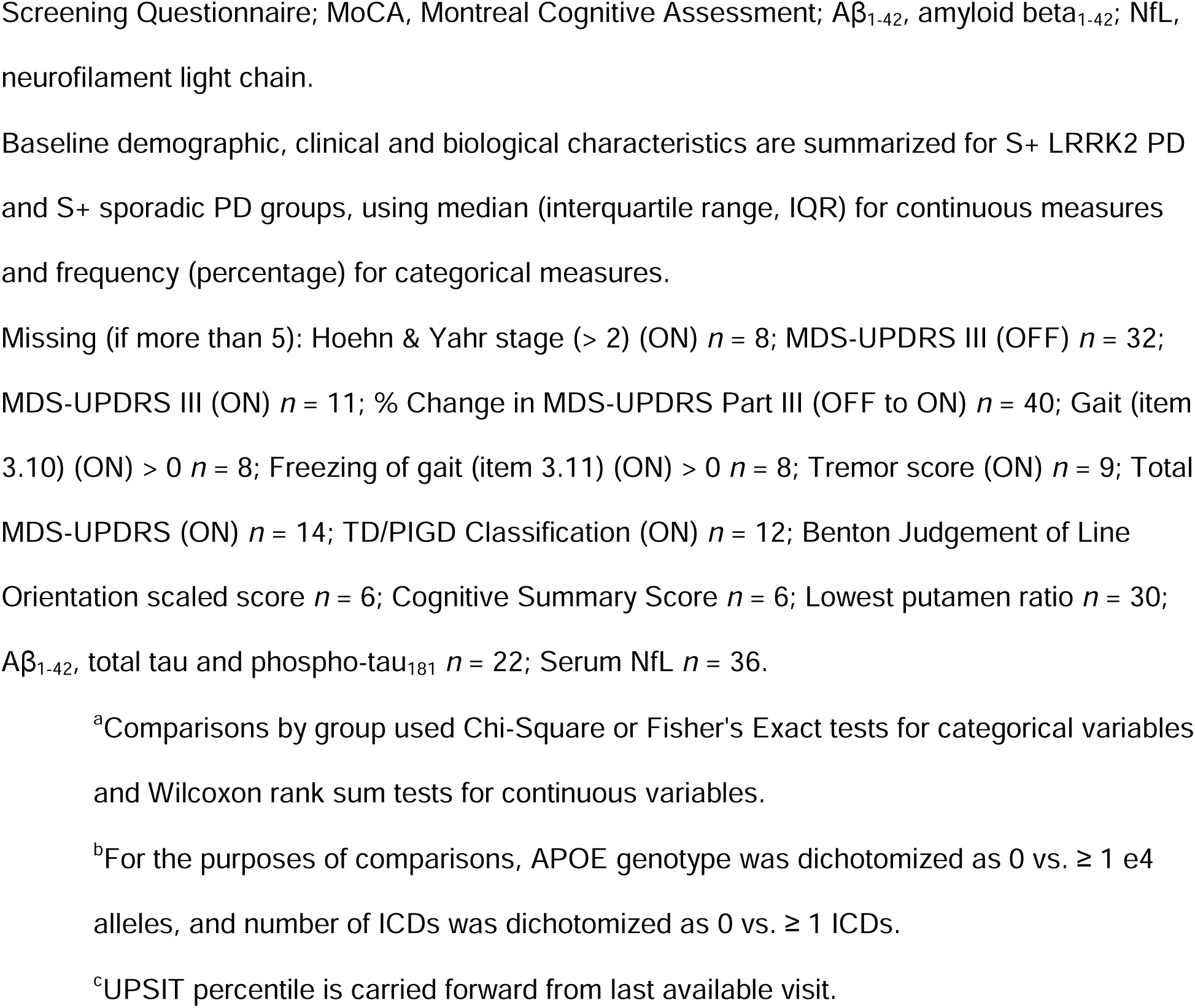
Demographics, clinical and biological characteristics of S+ LRRK2-PD and S+ sporadic PD participants.

### Longitudinal analysis

Raw mean and median values of clinical features and available biomarkers at each follow-up visit are detailed in Supplementary Table 3. Results of linear mixed effects models are shown in Supplementary Table 4. Progression slopes are presented in Figure 1. None of the clinical motor or non-motor measures showed differences in progression between the two groups aside from a marginal difference in the rate of change in the tremor score (ON) (p = 0.049), although neither LRRK2 or PD groups showed significant change over time (estimate [95% CI] −0.23 [−0.52, 0.06], p = 0.119; 0.16 [−0.12, 0.44], p = 0.257 respectively).

**Figure 1:**
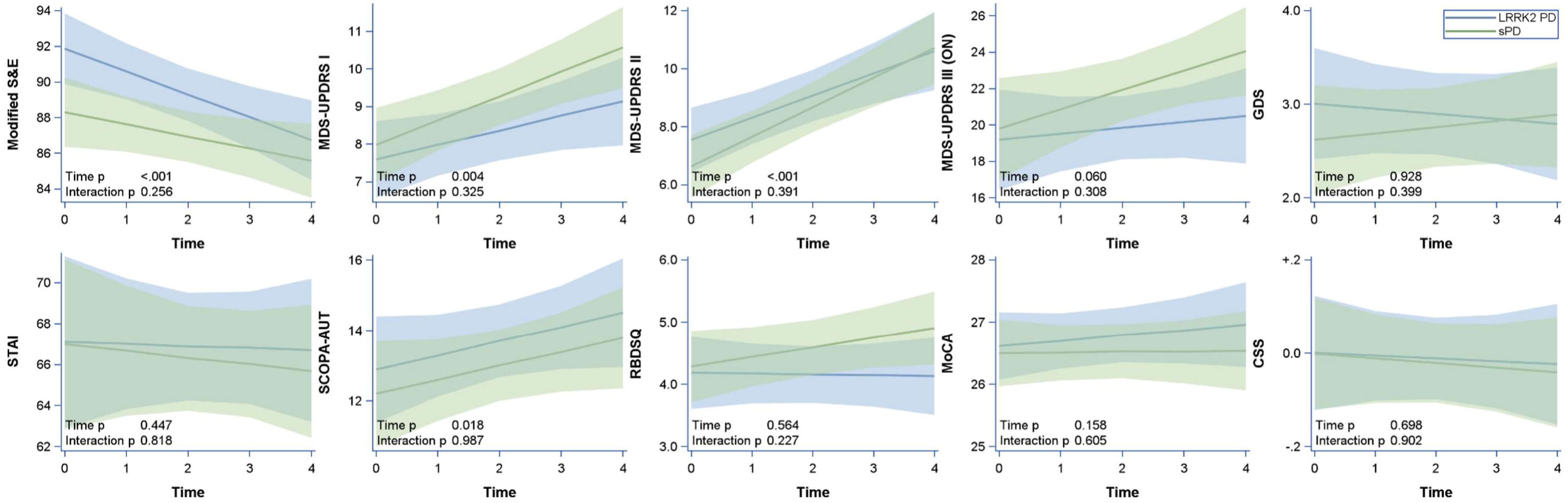
Predicted longitudinal trajectories (and 95% confidence bands) of selected clinical outcomes over four year follow up. Significance of linear mixed effects models indicated by overall time p-value and interaction (time x group) p-value, α = 0.05. Abbreviations: LRRK2 PD, alpha-synuclein positive LRRK2-Parkinson’s disease; sPD, alpha-synuclein positive sporadic Parkinson’s disease; Modified S&E, Modified Schwab and England Activities of Daily Living scale; MDS-UPDRS, Movement Disorder Society – Unified Parkinson’s Disease Rating Scale; GDS, Geriatric Depression Scale; STAI, State-Trait Anxiety Inventory; SCOPA-AUT, Scales for Outcomes in Parkinson’s disease – Autonomic Dysfunction; RBDSQ, REM Sleep Behavior Disorder Screening Questionnaire; MoCA, Montreal Cognitive Assessment; CSS, Cognitive Summary Score.

## Discussion

We report an analysis of S+ LRRK2-PD (predominantly G2019S) compared to S+ sPD with cohorts propensity matched by age, sex, disease duration and LED. Our data indicate that while S+ LRRK2-PD is characterized by milder motor clinical features and less DAT deficit at baseline, longitudinal trajectories do not differ significantly between the groups. These results, while requiring validation in other cohorts, have implications on our understanding of interaction between LRRK2 and αSyn-driven pathobiology in PD.

Historically, LRRK2-PD has been associated with milder phenotype and fewer non-motor features; however, these observations were made prior to the availability of *in vivo* biomarker of αSyn pathology.^8, 9^ LRRK2-PD is heterogeneous regarding the underlying proteinopathy even within same families, and in some cases is associated with pure nigrostriatal degeneration in absence of any protein aggregation.^3, 4^ Through this unique heterogeneity, LRRK2-PD serves as a valuable model through which the neurobiological underpinnings of clinical progression in PD—and in particular the role of αSyn—can be examined.

Our recently published data demonstrated that S- compared to S+ LRRK2-PD individuals have milder baseline phenotype and slower functional decline.^7^ The most striking differences included significant female sex predominance and lower rate of hyposmia.^7^ We did not elicit other biomarker separation between the groups aside from SAA status. These results suggest that previously reported milder LRRK2-PD phenotype may be driven by the S-subset of LRRK2-PD. The current analysis supports that hypothesis.

The whole discussion regarding the S-LRRK2-PD subset must be interpreted with caution considering recent reports of presence of oligomeric αSyn in Lewy body negative postmortem brains using proximity ligation assays.^18, 19^ If confirmed, the debate will refocus on understanding the implications not of binary αSyn status but of the protein conformation changes and potentially “the load” of pathology for which a quantitative *in vivo* marker is essential.

Several limitations to this study should be highlighted, particularly regarding sample selection, and genetic and ethnic features of the sample.

We used propensity score matching to establish groups matched for age, disease duration, sex, and LED. This approach was taken in an effort to address differences in baseline characteristics of cohorts specific to the PPMI dataset due to known differences in inclusion criteria for participants with sporadic versus genetic PD.^16, 17^ While it is recognized that matching procedures reduce the available sample size and can introduce bias, an approach without matching presented risk of comparing groups with differences reflective of recruitment as opposed to true neurobiological difference. Analyses of additional cohorts with a uniform recruitment strategy will be important in confirming our findings. We also acknowledge limitation in interpretation of motor score results using MDS-UPDRS III ON scores. While statistical analysis adjusted for LED, this similarly underscores the importance of analysis of other cohorts with diverse recruitment and data collection strategies.

Our analysis could be strengthened by comparison between S- LRRK2 PD and S+ sPD groups using the same methodology applied in this analysis. The sample size of the S-LRRK2 PPMI cohort was not adequate to support an analysis with a valid uniform matching technique and therefore was deferred. This comparison could be considered in different cohorts or with future growth of the LRRK2-PD PPMI cohort.

LRRK2-PD is genetically heterogeneous, with multiple associated causal variants including G2019S and R1441C/G along with other less frequently observed variants.^7, 10, 20^ Over 95% of S+ LRRK2 PPMI participants are p.G2019S carriers of European ancestry, while the S- group had greater diversity in variants, including R1441G among others.^7^ Given this difference, care must be taken in extrapolating the findings presented here acknowledging they reflect primarily features of G2019S LRRK2-PD and the effect of SAA status on progression of other variant carriers remains to be examined. This is of particular relevance given recent reports that the differences in motor decline between S+ and S- LRRK2-PD cohorts may be driven primarily by R1441C/G + M1646T carriers, a group that was not represented in the present analysis.^12^ We also acknowledge a difference in Hispanic ethnicity between groups which may introduce an additional source of potential bias. Further analysis of other LRRK2 variants in ethnically diverse cohorts is warranted.

## Conclusion

Although there are baseline features of LRRK2-PD that remain distinct from sPD even among the subset of participants with evidence of αSyn aggregation, the results of this analysis suggest that S+ LRRK2-PD behaves similarly to sPD in terms of clinical progression. This finding has important implications in ongoing efforts to understand biological drivers of PD progression. Furthermore, we emphasize that these findings make an argument for assessment of αSyn biomarkers in clinical trial design, enrollment, and analysis, including genetic cohorts with known proteinopathy heterogeneity like LRRK2-PD.

## Acknowledgement

We thank the PPMI – a public-private partnership – is funded by the Michael J. Fox Foundation for Parkinson’s Research and funding partners, including 4D Pharma, Abbvie, AcureX, Allergan, Amathus Therapeutics, Aligning Science Across Parkinson’s, AskBio, Avid Radiopharmaceuticals, BIAL, BioArctic, Biogen, Biohaven, BioLegend, BlueRock Therapeutics, Bristol-Myers Squibb, Calico Labs, Capsida Biotherapeutics, Celgene, Cerevel Therapeutics, Coave Therapeutics, DaCapo Brainscience, Denali, Edmond J. Safra Foundation, Eli Lilly, Gain Therapeutics, GE HealthCare, Genentech, GSK, Golub Capital, Handl Therapeutics, Insitro, Jazz Pharmaceuticals, Johnson & Johnson Innovative Medicine, Lundbeck, Merck, Meso Scale Discovery, Mission Therapeutics, Neurocrine Biosciences, Neuron23, Neuropore, Pfizer, Piramal, Prevail Therapeutics, Roche, Sanofi, Servier, Sun Pharma Advanced Research Company, Takeda, Teva, UCB, Vanqua Bio, Verily, Voyager Therapeutics, the Weston Family Foundation and Yumanity Therapeutics.

## Authors Roles

(1) Research Project: A. Conception, B. Organization, C. Execution; (2) Statistical Analysis: A. Design, B. Execution, C. Review and Critique; (3) Manuscript Preparation:

A. Writing of the First Draft, B. Review and Critique.

L.M 1B, 1C, 2A, 2C, 3A, 3B

S.H.C 1B, 1C, 2A, 2B, 2C, 3B

D.E.L 1B, 1C, 2A, 2B, 2C, 3B

C.G 1B, 1C, 2A, 2B, 2C, 3B

L.M.C 1A, 1C, 2A, 2C, 3B

K.M.M 1A, 2C, 3B

B. W 2C, 3B

T.S 1A, 1B, 1C, 2A, 2C, 3B

### Financial Disclosures of all authors (for the preceding 12 months)

L.M has received travel expense reimbursement from the Parkinson Study Group.

S.H.C. is funded by grants from the Michael J Fox Foundation for Parkinson’s Research.

D.E.L. is funded by grants from the Michael J Fox Foundation for Parkinson’s Research.

L.M.C. declares consulting fees and research support from the Michael J Fox Foundation.

K.M.M. declares consultancies for Axial Therapeutics, Asceneuron, Calico, Hanall, JnJ, Michael J. Fox Foundation, Nitrase Therapeutics, NuraBio, NRG Therapeutics, Rome Therapeutics, Schrodinger, Ventyx, private equity companies. Serves on the BoD for Envisagenics, Retromer Tx; serves on the SAB for Axial, Nitrase, NRG, Sinopia, Vanqua; Received research Funding support from Michael J. Fox Foundation and Honoraria from ASAP.

T.S. declares consultancies for AcureX, Adamas, AskBio, Amneal, Blue Rock Therapeutics, Critical Path for Parkinson’s Consortium, Denali, The Michael J. Fox Foundation, Neuroderm, Roche, Sanofi, Sinopia, Takeda, and Vanqua Bio; on advisory boards for AcureX, Adamas, AskBio, Biohaven, Denali, GAIN, Neuron23 and Roche; on scientific advisory boards for Koneksa, Neuroderm, Sanofi and UCB; and received research funding from Amneal, Biogen, Roche, Neuroderm, Sanofi, Prevail and UCB and an investigator for NINDS, MJFF, Parkinson’s Foundation.

B.W. is an employee of Berry Consultants, LLC, in which capacity she serves as a consultant to numerous pharmaceutical and device companies on topics of statistical modeling and trial design. The identities of these clients are protected under non-disclosure agreements. All payments are made to Berry Consultants, LLC.

### Ethical Compliance Statement

This study did not require review by institutional review board or ethics committee. Data used are publicly available as stated in the data availability statement. Each PPMI recruitment site received approval from an institutional review board or ethics committee on human experimentation before study initiation. Written informed consent for research was obtained from all individuals participating in the PPMI study. We confirm that we have read the journal’s position on issues involved in ethical publication and affirm that this work is consistent with those guidelines.

## Appendix 1: PPMI Study Committees, Cores, and Collaborators

### PPMI EXECUTIVE STEERING COMMITTEE

Kenneth Marek, MD^1^ (Principal Investigator); Tanya Simuni, MD^2^; Andrew Siderowf, MD^3^; Caroline Tanner, MD^4^; Thomas F Tropea, DO^1^; Tatiana Foroud, PhD^5^; Lana Chahine, MD^6^; Brit Mollenhauer, MD^7^; Kalpana Merchant, MD^2^; Douglas Galasko, MD^8^; Christopher Coffey, PhD ^9^; Kathleen Poston, MD^10^; Roseanne Dobkin, PhD^11^; Ethan Brown, MD^4^; Roy Alcalay, MD^12^; Dan Weintraub, MD^3^; Emily Flagg, BA^1^; Kimberly Fabrizio, BA^1^

### PPMI STEERING COMMITTEE

Susan Bressman, MD^13^; Cornelis Blauwendraat, PhD^14^; Paola Casalin, PhD^15^; Sonya Dumanis, PhD^14^; Raymond James, RN^16^; Karl Kieburtz, MD^17^; Sneha Mantri, MS^18^; Werner Poewe, MD^19^; Michael Schwarzschild, MD^20^; John Seibyl^1^, MD; David Standaert, PhD^21^; Duygu Tosun-Turgut, PhD^4^

### MICHAEL J. FOX FOUNDATION

Sohini Chowdhury, MA^22^; Jamie Eberling, PhD^22^; Mark Frasier, PhD^22^; Leslie Kirsch, EdD^22^; Katie Kopil, PhD^22^; Maggie Kuhl, BA^22^; Alyssa O’Grady, BA^22^; Todd Sherer, PhD^22^; Tawny Willson, MBS^22^

### PPMI STUDY CORES

Project Management Core: Emily Flagg, BA^1^

Site Management Core: Tanya Simuni, MD^2^; Bridget McMahon, BS^1^

Data Strategy and Technical Operations: Craig Stanley, PhD^1^; Kim Fabrizio, BA^1^

Data Management Core: Dixie Ecklund, MBA^9^, MSN; Christine Kohnen, PhD^9^

Screening Core: Tatiana Foroud, PhD^5^; Laura Heathers, BA^5^; Christopher Hobbick, BSCE^5^; Gena Antonopoulos, BSN^5^

Imaging Core: John Seibyl, MD^1^; Kathleen Poston, MD^10^

Statistics Core: Christopher Coffey, PhD ^9^; Chelsea Caspell-Garcia, MS ^9^; Michael Brumm, MS ^9^

Bioinformatics Core: Arthur Toga, PhD^23^; Karen Crawford, MLIS^23^

Biorepository Core: Tatiana Foroud, PhD^5^; Jan Hamer, BS^5^

Biologics Review Committee: Brit Mollenhauer, MD^7^; Doug Galasko, MD ^8^; Kalpana Merchant, MD^2^

Genetics Core: Andrew Singleton, PhD^24^

Pathology Core: Tatiana Foroud, PhD^5^; Dirk Keene, MD^5^

Found: Caroline Tanner, MD^4^; Ethan Brown, MD^4^

PPMI Online: Carlie Tanner, MD^4^; Ethan Brown, MD^4^; Lana Chahine, MD^6^; Roseann Dobkin, PhD^11^; Monica Korell, MPH^4^

### PPMI SITE INVESTIGATORS

Neha Prakash MD^1^; Tanya Simuni, MD^2^; Nabila Dahodwala MD^3^; Caroline Tanner, MD^4^; Lana Chahine MD^6^; Brit Mollenhauer MD^7^; Sebastian Schade MD^7^; Douglas Galasko, MD^8^; Anat Mirelman PhD^12^; Roy Alcalay MD^12^; Katherine Leaver MD^13^; Marie Saint-Hilaire MD^16^; Ruth Schneider MD^17^; Christopher Tarolli MD^17^; Werner Poewe, MD^19^; Aleksandar Videnovic MD^20^; David Standaert PhD^21^; Marissa Dean, MD^21^; Sonja Jonsdottir PhD^25^; Rejko Krueger MD^25^; Claire Pauly PhD^25^; Stewart Factor DO^26^; Penelope Hogarth MD^26^; Robert Hauser MD^28^; Amy Amara PhD^29^; Michelle Fullard MD^29^; Cyrus Zabetian MD^30^; Hubert Fernandez MD^31^; Kathrin Brockmann MD^32^; Isabel Wurster PhD^32^; Yen Tai PhD^33^; Paolo Barone PhD^34^; Marina Picillo MD^34^; Stuart Isaacson MD^35^; Alberto Espay MD^36^; Eduardo Tolosa PhD^37^; Javier Ruiz Martinez PhD^38^; Leonidas Stefanis PhD^39^; Kelvin Chou MD^40^; Lorraine Kalia MD^41^; Connie Marras PhD^41^; David Grimes MD^42^; Tiago Mestre PhD^42^; Rajesh Pahwa MD^43^; Mark Lew MD^44^; Holly Shill MD^45^; Shyamal Mehta MD^46^; Giulietta Riboldi MD^47^; Nikolaus McFarland PhD^48^; Ron Postuma MD^49^; Zoltan Mari MD^50^; David Ledingham MD^51^; Nicola Pavese PhD^51^; Michele Hu PhD^52^; Norbert Brueggemann MD^53^,; Christine Klein MD^53^; Bastiaan Bloem PhD^54^; Cristina Simonet PhD^55^; Alastair Noyce PhD^55^; Anette Janzen PhD^56^; David Pedrosa MD^56^; Wolfgang Oertel PhD^56^; Njideka Okubadejo MD^57^ David Shprecher DO^58^; Arjun Tarakad MD^59^; Emile Moukheiber MD^60^

### PPMI SITE COORDINATORS

Joy Antala^1^; Carla Aranda^2^; Karen Williams^2^; Sophia Melton^2^; Karina Benson^2^; Ashwini Ramachandran^3^, Danielle Potts^3^; Grace LaMoure^3^; Ritikha Vengadesh^3^; Ryan Manzler^3^; Jaime Heller^4^; Primi Ranola^4^; Farah Kausar^4^; Sherri Mosovsky^6^; Diana Willeke^7^; Elizabeth Kalinkara Gomez^7^; Janelle Rodriguez^8^; Nobuko Kemmotsu^8^; May Eshel^12^; Deborah Raymond^13^; Abigail Desrosiers^16^; Raymond James^16^; Lauren Jackson^17^; Iris Egner^19^; Wesley Schlett^20^; Courtney Blair^21^; Lauren Ruffrage^21^; Berenice Sevilla^25^; Barbara Sommerfeld^26^; Dustin Le^27^; Erica Botting^28^; Gabriella Mazur^28^; Daniele Derlein^29^; Evan Doll^29^; Ying Liu^29^; Ciera Cobb^30^; Olivia Masiewicz^30^; Jennifer Mule^31^; Michael Morsillo^31^; Ella Hilt^32^; Aldazier Jakiran^33^; Dominga Valentino^34^; Lisbeth Pennente^35^; Bobbie Stubbeman^36^; Alicia Garrido^37^; Valeria Ravasi^37^; Ioana Croitoru^38^; Christos Koros^39^; Nikolas Papagiannakis^39^; Frank Ferrari^40^; Mengyu Zheng^41^; Shawna Reddie^42^; Alicia Alejandra^43^; Andrea Gray^43^; Alejandra Valenzuela^44^; Caitlin Goodman^45^; Sara Dresler^46^; Neil Santos^46^; Fahrial Esha^47^; Kyle Rizer^48^; Nadine Zablith^49^; Liliana Dumitrescu^50^; Debra Galley^51^; Victoria Kate Foster^51^; Jamil Razzaque^52^; Madita Grümmer^53^; Yara Krasowski^54^; Natalie Donkor^55^; Elisabeth Sittig^56^; Oluwadamilola Ojo^57^; Kelly Clark^58^; Rory Mahabir^59^; Kori Ribb^60^; Shamera Willoughby^60^

#### Institutions and affiliations

1. Institute for Neurodegenerative Disorders; New Haven, CT, USA
2. Northwestern University; Evanston, IL, USA
3. University of Pennsylvania; Philadelphia, PA, USA
4. University of California, San Francisco; San Francisco, CA, USA
5. Indiana University; Indianapolis, IN, USA
6. University of Pittsburgh; Pittsburgh, PA, USA
7. Paracelsus-Elena Klinik; Kassel, Germany
8. University of California, San Diego, San Diego, CA, USA
9. University of Iowa; Iowa City, IA, USA
10. Stanford University; Stanford, CA, USA
11. Rutgers University; New Brunswick, NJ, USA
12. Tel Aviv Sourasky Medical Center; Tel Aviv, Israel
13. Mount Sinai Beth Israel; New York, NY, USA
14. Coalition for Aligning Science; Chevy Chase, MD, USA
15. BioRep; Milan, Italy
16. Boston University School of Medicine; Boston, MA, USA
17. University of Rochester; Rochester, NY, USA
18. Duke University; Durham, NC, USA
19. University of Innsbruck; Innsbruck, Austria
20. Massachusetts General Hospital; Boston, MA, USA
21. University of Alabama at Birmingham; Birmingham, AL, USA
22. The Michael J. Fox Foundation for Parkinson’s Research; New York, NY, USA
23. Laboratory of Neuroimaging (LONI), USC; Los Angeles, CA, USA
24. National Institute on Aging, NIH; Bethesda, MD, USA
25. University of Luxembourg; Esch-sur-Alzette, Luxembourg
26. Emory University; Atlanta, GA, USA
27. Oregon Health and Science University; Portland, OR, USA
28. University of South Florida; Tampa, FL, USA
29. University of Colorado; Aurora, CO, USA
30. VA Puget Sound Health System; Seattle, WA, USA
31. Cleveland Clinic; Cleveland, OH, USA
32. University of Tubingen; Tubingen, Germany
33. Imperial College of London; London, UK
34. University of Salerno; Salerno, Italy
35. Parkinson’s Disease and Movement Disorders Center; Boca Raton, FL, USA
36. University of Cincinnati; Cincinnati, OH, USA
37. Hospital Clinic of Barcelona; Barcelona, Spain
38. Hospital Universitario Donostia; San Sebastian, Spain
39. University of Athens; Athens, Greece
40. University of Michigan; Ann Arbor, MI, USA
41. Toronto Western Hospital; Toronto, Canada
42. The Ottawa Hospital; Ottawa, Canada
43. University of Kansas Medical Center; Kansas City, KS, USA
44. Keck School of Medicine of the University of Southern California; Los Angeles, CA, USA
45. Barrow Neurological Institute; Phoenix, AZ, USA
46. Mayo Clinic Arizona; Scottsdale, AZ, USA
47. NYU Langone Medical Center; New York, NY, USA
48. University of Florida; Gainesville, FL, USA
49. Montreal Neurological Institute and Hospital/McGill; Montreal, C, Canada
50. Cleveland Clinic-Las Vegas Lou Ruvo Center for Brain Health; Las Vegas, NV, USA
51. Clinical Ageing Research Unit; Newcastle, UK
52. John Radcliffe Hospital Oxford and Oxford University; Oxford, UK
53. University of Luebeck; Luebeck, Germany
54. Radboud University; Nijmegen, Netherlands
55. □ueen Mary University of London; London, UK
56. Philipps-University Marburg; Marburg, Germany
57. University of Lagos; Lagos, Nigeria
58. Banner Sun Health Research Institute; Sun City, AZ, USA
59. Baylor College of Medicine; Houston, TX, USA
60. Johns Hopkins University; Baltimore, MD, USA

## Figure and Supplemental File Legends

**Supplementary Figure 1**

Flowchart of participant selection for analysis population.

Abbreviations: PD, Parkinson’s disease; sPD, sporadic PD; S-, alpha-synuclein negative; S+, alpha-synuclein positive; MSA, multiple system atrophy; LED, levodopa equivalent daily dose

**Supplementary Table 1**

Abbreviations: S+; alpha-synuclein positive; PD, Parkinson’s disease; LED, levodopa equivalent dose (mg)

Note: Baseline was defined to be a participant’s first visit on treatment; LRRK2 and sporadic PD participants were required to have at least 1 and 3 years of follow-up post-treatment initiation, respectively, to be considered for matching.

^a^Comparisons by group used Chi-Square or Fisher’s Exact tests for categorical variables and Wilcoxon rank sum tests for continuous variables.

**Supplementary Table 2**

Abbreviations: S+; alpha-synuclein positive; PD, Parkinson’s disease; LED, levodopa equivalent dose.

^a^Comparisons by group used Chi-Square or Fisher’s Exact tests for categorical variables and Wilcoxon rank sum tests for continuous variables.
^b^For the purposes of comparisons, APOE genotype was dichotomized as 0 vs. ≥ 1 e4 alleles.

**Supplementary Table 3**

Abbreviations: S+; alpha-synuclein positive; PD, Parkinson’s disease; MDS-UPDRS, Movement Disorders Society—Modified Unified Parkinson’s Disease Rating Scale; SCOPA-AUT, Scales for Outcomes in Parkinson’s disease–Autonomic Dysfunction; RBDSQ, REM Sleep Behavior Disorder Screening Questionnaire; MoCA, Montreal Cognitive Assessment; Aβ_1-42_, amyloid beta_1-42_; NfL, neurofilament light chain.

*Note:* In cases where variables are missing less than 10% of values at all visits, the missing values are not shown.

**Supplementary Table 4**

Abbreviations: S+; alpha-synuclein positive; PD, Parkinson’s disease; MDS-UPDRS, Movement Disorders Society—Modified Unified Parkinson’s Disease Rating Scale; SCOPA-AUT, Scales for Outcomes in Parkinson’s disease–Autonomic Dysfunction; RBDSQ, REM Sleep Behavior Disorder Screening Questionnaire; MoCA, Montreal Cognitive Assessment.

Note: Models were adjusted for levodopa equivalent dose at each visit. In cases where the model did not converge, the relevant fields are left blank.

^a^For binary outcomes, the time effect is reported as odds ratio (95% CI for OR).

