## Supplementary tables for "Clinical progression in alpha-synuclein positive LRRK2-PD and sporadic Parkinson’s disease: a longitudinal analysis"

**Supplementary Table 1. Sample demographics and other characteristics of S+ LRRK2-PD and S+ sporadic PD participants prior to matching.**

| **Variable** | **LRRK2-PD (N = 96)** | **Sporadic PD (N = 284)** | ***p*-value^a^** |
| --- | --- | --- | --- |
| **Age at baseline, years**, median (IQR) | 61.5 [54.9-66.8] | 63.7 [56.5-70.2] | 0.019 |
| **Male sex**, n (%) | 59 (61%) | 191 (67%) | 0.301 |
| **Years since PD diagnosis**, median (IQR) | 2.3 [1.2-4.2] | 1.7 [1.3-2.6] | 0.053 |
| **LED**, median (IQR) | 500.0 [300.0-705.0] | 246.2 [120.0-346.2] | <.001 |

### **Supplementary Table 2. Sample demographics and other characteristics of S+ LRRK2-PD participants at baseline and S+ sporadic PD participants at year 2 prior to matching.**

| **Variable** | **LRRK2 PD (N = 98)** | **Sporadic PD (N = 279)** | ***p*-value^a^** |
| --- | --- | --- | --- |
| **Age at baseline, years**, median (IQR) | 61.5 [54.7-66.9] | 64.2 [56.8-70.9] | 0.006 |
| **Age at PD symptom onset, years**, median (IQR) | 57.2 [47.8-62.1] | 60.1 [52.9-66.7] | 0.002 |
| Missing | 4 | 6 |  |
| **Male sex**, n (%) | 59 (60%) | 187 (67%) | 0.222 |
| **Years of education**, median (IQR) | 17.0 [14.0-19.0] | 16.0 [14.0-18.0] | 0.008 |
| **Years since PD diagnosis**, median (IQR) | 2.1 [1.1-4.0] | 2.4 [2.2-2.7] | 0.012 |
| **Race (% White)**, n (%) | 93 (95%) | 262 (94%) | 0.809 |
| Missing | 0 | 1 |  |
| **Hispanic**, n (%) | 14 (14%) | 7 (3%) | <.001 |
| **LED**, median (IQR) | 450.0 [250.0-700.0] | 300.0 [100.0-450.0] | <.001 |
| **LRRK2 variant**, n (%) |  |  |  |
| G2019S | 93 (95%) | - |  |
| R1441G | 4 (4%) | - |  |
| R1441C | 1 (1%) | - |  |
| **APOE genotype - number of e4 alleles^b^**, n (%) |  |  | 0.655 |
| 0 | 78 (80%) | 216 (77%) |  |
| 1 | 19 (19%) | 57 (20%) |  |
| 2 | 1 (1%) | 6 (2%) |  |

### **Supplementary Table 3. Longitudinal assessment of motor, non-motor, imaging and biofluid biomarkers of matched S+ LRRK2-PD and S+ sporadic PD participants.**

| **Variable** | **Baseline** | **Year 1** | **Year 2** | **Year 3** | **Year 4** |
| --- | --- | --- | --- | --- | --- |
|  | **LRRK2 N=79 sPD N=79** | **LRRK2 N=76 sPD N=76** | **LRRK2 N=62 sPD N=73** | **LRRK2 N=61 sPD N=74** | **LRRK2 N=59 sPD N=65** |
| **Modified Schwab and England**, mean (SD) | | | | | |
| LRRK2 PD | 92 (8) | 92 (8) | 91 (10) | 89 (10) | 89 (10) |
| sPD | 88 (7) | 86 (9) | 86 (9) | 85 (9) | 85 (10) |
| **Hoehn & Yahr stage (> 2) (ON)**, n (%) | | | | | |
| LRRK2 PD | 0 (0%) | 1 (1%) | 0 (0%) | 3 (5%) | 3 (5%) |
| Missing | 4 | 2 | 5 | 1 | 0 |
| sPD | 1 (1%) | 4 (6%) | 6 (8%) | 7 (10%) | 5 (8%) |
| Missing | 4 | 11 | 0 | 1 | 2 |
| **MDS-UPDRS I**, mean (SD) | | | | | |
| LRRK2 PD | 7 (5) | 8 (5) | 8 (4) | 9 (5) | 9 (5) |
| sPD | 7 (5) | 9 (6) | 9 (6) | 10 (7) | 10 (6) |
| **MDS-UPDRS II**, mean (SD) | | | | | |
| LRRK2 PD | 7 (4) | 8 (5) | 8 (5) | 9 (6) | 9 (6) |
| sPD | 8 (5) | 8 (6) | 9 (6) | 11 (7) | 11 (7) |
| **MDS-UPDRS III (ON)**, mean (SD) | | | | | |
| LRRK2 PD | 18 (10) | 17 (10) | 19 (10) | 19 (10) | 18 (9) |
| Missing | 5 | 2 | 7 | 3 | 6 |
| sPD | 23 (12) | 23 (14) | 24 (15) | 25 (14) | 26 (13) |
| Missing | 6 | 11 | 0 | 1 | 2 |
| **Gait (item 3.10) (ON) > 0**, n (%) | | | | | |
| LRRK2 PD | 45 (60%) | 47 (64%) | 35 (61%) | 35 (58%) | 35 (59%) |
| Missing | 4 | 2 | 5 | 1 | 0 |
| sPD | 51 (68%) | 43 (66%) | 49 (67%) | 53 (73%) | 47 (75%) |
| Missing | 4 | 11 | 0 | 1 | 2 |
| **Freezing of gait (item 3.11) (ON) > 0**, n (%) | | | | | |
| LRRK2 PD | 3 (4%) | 2 (3%) | 3 (5%) | 2 (3%) | 2 (3%) |
| Missing | 4 | 2 | 5 | 1 | 0 |
| sPD | 2 (3%) | 1 (2%) | 1 (1%) | 5 (7%) | 2 (3%) |
| Missing | 4 | 11 | 0 | 1 | 2 |
| **Tremor score (ON)**, mean (SD) | | | | | |
| LRRK2 PD | 3 (4) | 3 (3) | 3 (3) | 3 (3) | 2 (3) |
| Missing | 5 | 2 | 5 | 1 | 1 |
| sPD | 4 (4) | 3 (3) | 3 (3) | 4 (4) | 4 (4) |
| Missing | 4 | 11 | 0 | 1 | 2 |
| **Total MDS-UPDRS (ON)**, mean (SD) | | | | | |
| LRRK2 PD | 31 (15) | 33 (15) | 34 (15) | 36 (16) | 35 (14) |
| Missing | 7 | 3 | 7 | 3 | 6 |
| sPD | 38 (17) | 39 (21) | 42 (21) | 46 (21) | 47 (20) |
| Missing | 7 | 11 | 0 | 1 | 2 |
| **Geriatric Depression Scale**, mean (SD) | | | | | |
| LRRK2 PD | 3 (3) | 3 (3) | 3 (3) | 3 (3) | 3 (3) |
| sPD | 2 (3) | 3 (3) | 2 (2) | 3 (3) | 3 (3) |
| **State-Trait Anxiety Inventory**, mean (SD) | | | | | |
| LRRK2 PD | 67 (18) | 68 (20) | 67 (17) | 69 (21) | 67 (20) |
| sPD | 64 (17) | 66 (18) | 65 (18) | 65 (19) | 64 (18) |
| **SCOPA-AUT**, mean (SD) | | | | | |
| LRRK2 PD | 12 (7) | 13 (8) | 14 (8) | 14 (9) | 14 (8) |
| sPD | 11 (7) | 13 (8) | 13 (8) | 14 (8) | 14 (8) |
| **RBDSQ**, mean (SD) | | | | | |
| LRRK2 PD | 4 (2) | 4 (3) | 4 (3) | 4 (3) | 4 (3) |
| sPD | 4 (3) | 5 (3) | 5 (3) | 5 (3) | 5 (3) |
| **RBDSQ ≥ 6**, n (%) | | | | | |
| LRRK2 PD | 14 (18%) | 17 (22%) | 13 (22%) | 11 (18%) | 17 (29%) |
| sPD | 25 (32%) | 27 (36%) | 26 (36%) | 29 (39%) | 28 (43%) |
| **Epworth Sleepiness Scale**, mean (SD) | | | | | |
| LRRK2 PD | 7 (4) | 8 (4) | 8 (5) | 7 (5) | 8 (4) |
| sPD | 8 (4) | 7 (4) | 8 (5) | 8 (5) | 9 (5) |
| **MoCA**, mean (SD) | | | | | |
| LRRK2 PD | 27 (3) | 27 (3) | 27 (2) | 27 (2) | 27 (2) |
| Missing | 0 | 0 | 5 | 3 | 6 |
| sPD | 26 (3) | 26 (3) | 26 (3) | 26 (5) | 27 (3) |
| Missing | 2 | 2 | 1 | 0 | 1 |
| **Hopkins Verbal Learning Test Delayed Recall t-score**, mean (SD) | | | | | |
| LRRK2 PD | 46.3 (12.1) | 46.9 (11.9) | 48.0 (12.2) | 45.4 (13.0) | 46.0 (12.5) |
| Missing | 1 | 2 | 5 | 3 | 7 |
| sPD | 45.6 (11.8) | 44.3 (12.2) | 44.6 (13.5) | 45.1 (13.8) | 46.4 (13.0) |
| Missing | 3 | 1 | 1 | 1 | 3 |
| **Hopkins Verbal Learning Test Immediate/Total Recall t-score**, mean (SD) | | | | | |
| LRRK2 PD | 46.4 (11.1) | 47.7 (11.7) | 49.5 (10.3) | 45.7 (11.7) | 47.6 (11.0) |
| sPD | 43.7 (10.4) | 45.1 (12.9) | 46.4 (13.5) | 45.8 (12.9) | 47.1 (12.4) |
| **Benton Judgement of Line Orientation scaled score**, mean (SD) | | | | | |
| LRRK2 PD | 11.6 (3.2) | 11.1 (3.1) | 12.1 (2.3) | 10.9 (3.2) | 11.7 (3.0) |
| sPD | 12.2 (3.1) | 12.5 (2.9) | 11.9 (3.3) | 12.1 (3.1) | 12.0 (3.2) |
| **Letter Number Sequencing scaled score**, mean (SD) | | | | | |
| LRRK2 PD | 11.1 (2.9) | 11.0 (2.8) | 11.6 (2.8) | 11.2 (2.2) | 11.1 (2.6) |
| Missing | 2 | 2 | 5 | 4 | 7 |
| sPD | 11.0 (2.6) | 11.2 (2.8) | 10.8 (3.1) | 11.2 (3.3) | 11.3 (3.1) |
| Missing | 3 | 1 | 1 | 1 | 3 |
| **Symbol Digit Modalities Test t-score**, mean (SD) | | | | | |
| LRRK2 PD | 45.1 (9.8) | 46.6 (9.2) | 47.4 (9.4) | 48.4 (10.0) | 46.5 (8.5) |
| Missing | 1 | 1 | 5 | 3 | 6 |
| sPD | 44.7 (9.9) | 45.1 (11.8) | 45.1 (11.1) | 43.2 (11.2) | 44.9 (11.4) |
| Missing | 3 | 1 | 1 | 1 | 3 |
| **Semantic Fluency (Animals) t-score**, mean (SD) | | | | | |
| LRRK2 PD | 51.7 (12.1) | 53.9 (11.7) | 50.5 (10.8) | 52.1 (11.8) | 52.0 (12.8) |
| sPD | 50.3 (11.0) | 51.5 (10.9) | 50.2 (11.7) | 49.7 (11.7) | 51.6 (11.7) |
| **Cognitive Summary Score**, mean (SD) | | | | | |
| LRRK2 PD | -0.0 (0.7) | 0.0 (0.7) | 0.1 (0.6) | -0.0 (0.7) | 0.0 (0.6) |
| Missing | 3 | 2 | 5 | 4 | 8 |
| sPD | -0.1 (0.7) | -0.0 (0.8) | -0.1 (0.8) | -0.1 (0.8) | 0.0 (0.8) |
| Missing | 3 | 2 | 1 | 1 | 3 |
| **Number of Impulse Control Disorders ≥ 1**, n (%) | | | | | |
| LRRK2 PD | 28 (35%) | 33 (43%) | 20 (34%) | 22 (37%) | 17 (29%) |
| sPD | 18 (23%) | 19 (25%) | 15 (21%) | 23 (31%) | 18 (28%) |
| **Lowest putamen ratio**, mean (SD) | | | | | |
| LRRK2 PD | 0.30 (0.10) | 0.26 (0.04) | 0.25 (0.10) | 0.27 (0.02) | 0.22 (0.08) |
| Missing | 11 | 69 | 21 | 58 | 27 |
| sPD | 0.26 (0.08) | 0.25 (0.08) | 0.20 (0.09) | 0.22 (0.07) | 0.27 (N/A) |
| Missing | 19 | 30 | 57 | 38 | 64 |
| **Aβ_1-42_ (pg/mL)**, median [IQR] | | | | | |
| LRRK2 PD | 793.4 [639.9-1,030.7] | 875.0 [641.8-1,115.3] | 888.5 [616.7-1,121.4] | 795.0 [598.0-1,068.7] | 900.8 [560.3-1,237.3] |
| Missing | 8 | 24 | 25 | 33 | 40 |
| sPD | 814.8 [622.3-1,137.0] | 703.0 [540.5-1,107.0] | 768.0 [643.0-1,187.6] | 757.8 [586.9-1,260.9] | 608.5 [500.3-882.4] |
| Missing | 14 | 23 | 28 | 31 | 38 |
| **Aβ_1-42_ ≤ 683 pg/mL**, n (%) | | | | | |
| LRRK2 PD | 22 (31%) | 14 (27%) | 11 (30%) | 11 (39%) | 5 (26%) |
| Missing | 8 | 24 | 25 | 33 | 40 |
| sPD | 20 (31%) | 24 (45%) | 14 (31%) | 18 (42%) | 14 (52%) |
| Missing | 14 | 23 | 28 | 31 | 38 |
| **Aβ_1-42_ ≤ 710 pg/mL**, n (%) | | | | | |
| LRRK2 PD | 26 (37%) | 14 (27%) | 13 (35%) | 11 (39%) | 5 (26%) |
| Missing | 8 | 24 | 25 | 33 | 40 |
| sPD | 23 (35%) | 27 (51%) | 17 (38%) | 19 (44%) | 14 (52%) |
| Missing | 14 | 23 | 28 | 31 | 38 |
| **Total tau (pg/mL)**, median [IQR] | | | | | |
| LRRK2 PD | 140.1 [118.6-193.6] | 152.9 [119.7-186.4] | 140.1 [114.2-190.9] | 133.7 [107.9-176.3] | 138.9 [124.0-179.8] |
| Missing | 8 | 24 | 25 | 33 | 40 |
| sPD | 155.6 [122.7-210.2] | 147.3 [117.2-191.2] | 162.7 [132.6-208.0] | 156.5 [112.2-225.0] | 138.3 [106.9-162.0] |
| Missing | 14 | 22 | 27 | 31 | 38 |
| **Total tau ≥ 266 pg/mL**, n (%) | | | | | |
| LRRK2 PD | 4 (6%) | 6 (12%) | 3 (8%) | 1 (4%) | 1 (5%) |
| Missing | 8 | 24 | 25 | 33 | 40 |
| sPD | 4 (6%) | 3 (6%) | 4 (9%) | 4 (9%) | 2 (7%) |
| Missing | 14 | 22 | 27 | 31 | 38 |
| **Total tau ≥ 112 pg/mL**, n (%) | | | | | |
| LRRK2 PD | 60 (85%) | 46 (88%) | 28 (76%) | 20 (71%) | 18 (95%) |
| Missing | 8 | 24 | 25 | 33 | 40 |
| sPD | 53 (82%) | 42 (78%) | 38 (83%) | 34 (79%) | 20 (74%) |
| Missing | 14 | 22 | 27 | 31 | 38 |
| **Phospho-tau_181_ (pg/mL)**, median [IQR] | | | | | |
| LRRK2 PD | 12.3 [10.0-15.5] | 13.1 [10.0-16.3] | 13.9 [10.5-16.9] | 11.5 [9.4-14.5] | 13.9 [10.8-14.8] |
| Missing | 8 | 24 | 25 | 33 | 40 |
| sPD | 12.5 [9.8-17.3] | 12.0 [9.4-16.1] | 13.1 [10.2-17.5] | 12.2 [9.4-18.5] | 11.3 [9.3-13.6] |
| Missing | 14 | 22 | 27 | 31 | 38 |
| **Phospho-tau_181_ ≥ 24 pg/mL**, n (%) | | | | | |
| LRRK2 PD | 4 (6%) | 4 (8%) | 3 (8%) | 1 (4%) | 1 (5%) |
| Missing | 8 | 24 | 25 | 33 | 40 |
| sPD | 6 (9%) | 3 (6%) | 5 (11%) | 3 (7%) | 2 (7%) |
| Missing | 14 | 22 | 27 | 31 | 38 |
| **Phospho-tau_181_ ≥ 17.6 pg/mL**, n (%) | | | | | |
| LRRK2 PD | 12 (17%) | 12 (23%) | 8 (22%) | 5 (18%) | 3 (16%) |
| Missing | 8 | 24 | 25 | 33 | 40 |
| sPD | 15 (23%) | 13 (24%) | 10 (22%) | 11 (26%) | 5 (19%) |
| Missing | 14 | 22 | 27 | 31 | 38 |
| **Serum NfL (pg/mL)**, mean (SD) | | | | | |
| LRRK2 PD | 11.6 (4.2) | 14.3 (7.6) | 13.3 (5.1) | 13.3 (5.5) | 24.0 (3.1) |
| Missing | 20 | 20 | 22 | 30 | 56 |
| sPD | 13.6 (6.1) | 15.0 (7.8) | 15.1 (7.7) | 21.4 (23.6) | 18.6 (11.9) |
| Missing | 16 | 17 | 31 | 52 | 27 |

### **Supplementary Table 4. Results of linear mixed effects models in matched S+ LRRK2-PD and S+ sporadic PD participants.**

|  | **Linear Model^a^** | | | **Quadratic Model** | | **3-Way Interaction Model** | |
| --- | --- | --- | --- | --- | --- | --- | --- |
| **Variable** | **Interaction p-value** | **Time Effect (95% CI)** | **Time p-value** | **p-value** | **Effect** | **p-value** | **Effect** |
| Modified Schwab and England | 0.256 | -1.111 (-1.673, -0.550) | <0.001 | 0.591 | 0.264 | 0.219 | -1.399 |
| Hoehn & Yahr stage (> 2) (ON)^a^ |  |  |  |  |  |  |  |
| MDS-UPDRS I | 0.325 | 0.423 (0.140, 0.706) | 0.004 | 0.225 | 0.300 | 0.193 | 0.750 |
| MDS-UPDRS II | 0.391 | 0.890 (0.575, 1.205) | <0.001 | 0.702 | 0.105 | 0.509 | 0.414 |
| MDS-UPDRS III (ON) | 0.308 | 0.730 (-0.030, 1.489) | 0.060 | 0.125 | -0.897 | 0.675 | -0.655 |
| Gait (item 3.10) (ON) > 0^a^ | 0.105 | 1.001 (0.800, 1.252) | 0.993 | 0.955 | 0.013 | 0.275 | 0.508 |
| Freezing of gait (item 3.11) (ON) > 0^a^ | 0.436 | 1.221 (0.722, 2.064) | 0.455 | 0.448 | 0.454 | 0.592 | 0.840 |
| Tremor score (ON) | 0.049 | LRRK2: -0.231 (-0.521, 0.060) sPD: 0.161 (-0.118, 0.441) | LRRK2: 0.119 sPD: 0.257 | 0.122 | -0.279 | 0.430 | 0.337 |
| Total MDS-UPDRS (ON) | 0.069 | 2.040 (0.980, 3.099) | <0.001 | 0.483 | -0.574 | 0.556 | 1.283 |
| Geriatric Depression Scale | 0.399 | 0.007 (-0.147, 0.161) | 0.928 | 0.551 | -0.081 | 0.364 | -0.281 |
| State-Trait Anxiety Inventory | 0.818 | -0.404 (-1.450, 0.643) | 0.447 | 0.965 | 0.039 | 0.307 | -2.179 |
| SCOPA-AUT | 0.987 | 0.500 (0.085, 0.914) | 0.018 | 0.425 | -0.260 | 0.821 | -0.191 |
| RBDSQ | 0.227 | 0.044 (-0.106, 0.193) | 0.564 | 0.517 | 0.088 | 0.700 | -0.115 |
| RBDSQ ≥ 6^a^ | 0.863 | 1.108 (0.874, 1.406) | 0.396 | 0.330 | 0.250 | 0.264 | -0.755 |
| Epworth Sleepiness Scale | 0.416 | 0.332 (0.103, 0.561) | 0.005 | 0.115 | 0.358 | 0.222 | 0.559 |
| MoCA | 0.605 | 0.114 (-0.045, 0.272) | 0.158 | 0.523 | -0.089 | 0.644 | 0.149 |
| Hopkins Verbal Learning Test Delayed Recall t-score | 0.120 | -0.033 (-0.829, 0.763) | 0.935 | 0.821 | -0.169 | 0.293 | 1.706 |
| Hopkins Verbal Learning Test Immediate/Total Recall t-score | 0.414 | 0.181 (-0.551, 0.912) | 0.626 | 0.494 | 0.475 | 0.485 | 1.025 |
| Benton Judgement of Line Orientation scaled score | 0.083 | -0.058 (-0.240, 0.123) | 0.526 | 0.352 | -0.178 | 0.886 | -0.052 |
| Letter Number Sequencing scaled score | 0.555 | 0.003 (-0.152, 0.158) | 0.967 | 0.108 | -0.243 | 0.556 | -0.182 |
| Symbol Digit Modalities Test t-score | 0.358 | -0.117 (-0.629, 0.395) | 0.653 | 0.104 | -0.779 | 0.793 | -0.270 |
| Semantic Fluency (Animals) t-score | 0.806 | -0.223 (-0.854, 0.407) | 0.484 | 0.787 | 0.166 | 0.662 | 0.542 |
| Cognitive Summary Score | 0.902 | -0.007 (-0.040, 0.027) | 0.698 | 0.467 | -0.022 | 0.830 | 0.014 |
| Number of Impulse Control Disorders ≥ 1^a^ | 0.095 | 0.947 (0.771, 1.163) | 0.602 | 0.974 | 0.007 | 0.495 | -0.301 |
