## Supplementary figures and images for "Clinical progression in alpha-synuclein positive LRRK2-PD and sporadic Parkinson’s disease: a longitudinal analysis"

### Supplementary Figure 1

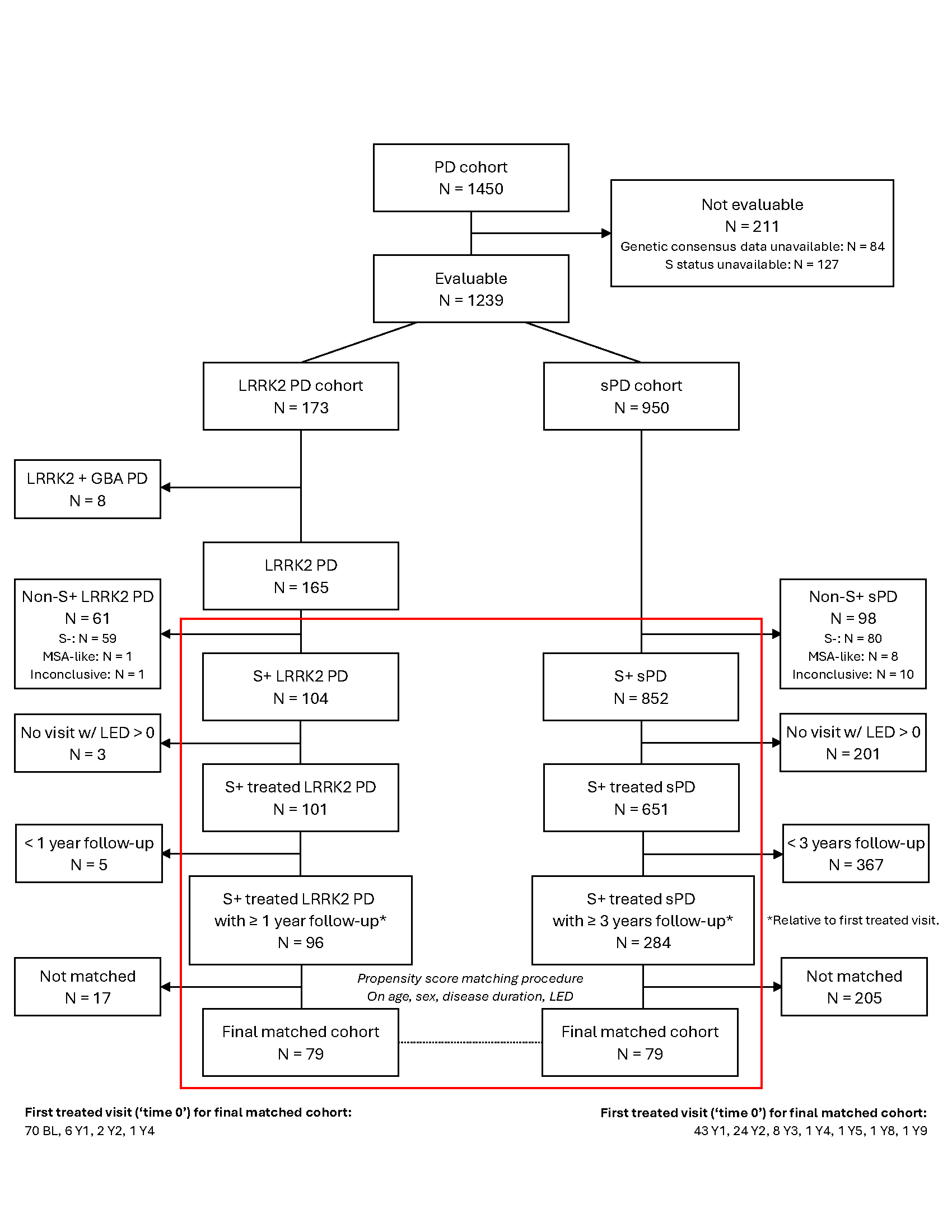
